# Introducing riskCommunicator: an R package to obtain interpretable effect estimates for public health

**DOI:** 10.1101/2022.03.02.22271755

**Authors:** Jessica A. Grembi, Elizabeth T. Rogawski McQuade

## Abstract

Common statistical modeling methods do not necessarily produce the most relevant or interpretable effect estimates to communicate risk. Overreliance on the odds ratio and relative effect measures limit the potential impact of epidemiologic and public health research. We created a straightforward R package, called `riskCommunicator`, to facilitate the presentation of a variety of effect measures, including risk differences and ratios, number needed to treat, incidence rate differences and ratios, and mean differences. The `riskCommunicator` package uses g-computation with parametric regression models and bootstrapping for confidence intervals to estimate effect measures in time-fixed data. We demonstrate the utility of the package using data from the Framingham Heart Study to estimate the effect of prevalent diabetes on the 24-year risk of cardiovascular disease or death.

The absolute 24-year risk of cardiovascular disease or death was 30% (95% confidence interval (CI): 22, 38) higher among subjects with diabetes compared to subjects without diabetes at baseline. The relative 24-year risk was 55% (95% CI: 40, 70) higher. Because the outcome was common (41.8%), the odds ratio (4.55) is highly inflated compared to the risk ratio (1.55). An expected 4 additional persons would need to have diabetes at baseline to observe an increase in the number of cases of cardiovascular disease or death by 1 over 24 years of follow-up.

The package promotes the communication of public-health relevant effects and is accessible to a broad range of epidemiologists and health researchers with little to no expertise in causal inference methods or advanced coding.

## Background

The communication of disease risk and the effects of exposures and interventions on that risk are core components of public health research and practice. Unfortunately, reporting of results from epidemiologic studies both in the published scientific literature and to the public is often confused by imprecise language, jargon, and incomplete reporting [1,2]. While it may be easiest to rely on the default output from standard functions in statistical programs, common statistical methods estimate parameters that are often not the most informative. Epidemiologists and the larger community of public health practitioners could benefit from easy-to-use tools to facilitate the presentation of relevant effects.

Overreliance on the odds ratio [3–6] and more broadly on relative effect measures [7,8] are two examples of opportunities to improve the reporting and interpretability of epidemiologic results. Efforts to increase the reporting of difference effect measures and risk ratios over odds ratios are not new, and several solutions have been previously proposed, including log-binomial and log-linear regression models, Poisson regression to approximate log-binomial regression when the latter does not converge [9], standardization-based approaches [10], linear-expit regression [11], and ordinary least-squares regression with transformed variables [12]. However, these models are not as efficient as logistic regression, can have convergence problems, and may require robust variance estimators [9,13].

Parametric g-computation is an attractive alternative because of the flexibility to estimate a variety of effect measures while relying on the preferable statistical properties of logistic regression for the parametric modeling. G-computation is conceptually equivalent to standardization, and the use of parametric models allows for highly-dimensional data and continuous covariates. G-computation has been applied to estimate risk differences and risk ratios from logistic regression models previously [14–16].

Despite the availability of g-computation-based methods, these methods are rarely used to estimate risk differences and risk ratios in standard time-fixed study designs. Recent applications of these methods have focused on complicated study designs, such as with longitudinal data with time-varying confounding affected by prior exposure [17]. In these applications, the methods are complex and difficult to understand and/or implement for the average data analyst. Coding requirements and computational limitations may also dissuade users from attempting these methods. Recently available R packages [18,19] and Statistical Analysis System (SAS) macros [20] are geared towards estimating these more complicated effects and may be overwhelming to new users.

We aimed to create a straightforward R package, called `riskCommunicator`, to facilitate the presentation of a variety of effect measures, including risk differences and ratios, number needed to treat, incidence rate differences and ratios, and mean differences, using g-computation. To make the package accessible to a broad range of health researchers, our goal was to design functions that were as easy to use as the standard logistic regression functions in R (e.g. `glm`) and that would require little to no expertise in causal inference methods or advanced coding.

## Implementation

The `riskCommunicator` package uses g-computation [21,22,16,17] with standard parametric regression models and bootstrapping for confidence intervals to estimate effect measures in the context of time-fixed exposure and outcome data. Broadly, the effects estimated are average treatment effects (ATEs), estimated for difference measures with a binary exposure variable as:

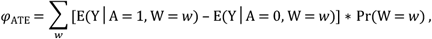

where Y is the outcome of interest, A is the exposure of interest, and W are covariates. In this way, the effects are standardized to the joint distribution of covariates in the total study population.

The package contains two main functions available to end users: `gComp` (the primary function) and `pointEstimate` (used internally within the `gComp` function, but provided to users in case of complex dependencies among observations, e.g. nested clusters-within-clusters, where a single cluster-level bootstrap resampling might not be optimal). `pointEstimate` computes a point estimate by executing three steps of g-computation. First, a regression of the outcome on the exposure and relevant covariates is fit using the provided dataset with a generalized linear model. The underlying model distribution is based on the outcome type supplied by the user (see `outcome.type` in Table 1 for details). Next, using the estimated parameters from the model, counterfactuals are predicted for each observation in the data set under each level of the exposure. Finally, the mean predicted value for each exposure regime across all observations is calculated and used to estimate marginal difference and ratio effects. The `gComp` function first estimates effects in the original data (using the `pointEstimate` function). Then, bootstrap resampling of the original dataset is conducted, and the `pointEstimate` function is called on each resample in order to estimate accurate standard errors and provide a 95% confidence interval (CI). Confidence intervals are based on the 2.5^th^ and 97.5^th^ percentiles of the bootstrap resampling results [23].

**Table 1.** Arguments supplied to the gComp function in the riskCommunicator package.

| Argument | Description |
| --- | --- |
| `data` | (Required) A `data.frame` or tibble containing variables for Y, X, and Z or with variables matching the model variables specified in a user-supplied formula. Data set should also contain variables for the optional `subgroup` and `offset`, if they are specified. |
| outcome.type | (Required) Character argument to describe the outcome type. Acceptable responses, and the corresponding error distribution and link function used in the `glm`, include:<br>**binary**<br>(Default) A binomial distribution with link = 'logit' is used.<br>**count**<br>A Poisson distribution with link = 'log' is used.<br>**count_nb**<br>A negative binomial distribution with link = 'log' is used, where the theta parameter is estimated internally; ideal for over-dispersed count data.<br>**rate**<br>A Poisson distribution with link = 'log' is used; ideal for events/person-time outcomes.<br>**rate_nb**<br>A negative binomial distribution with link = 'log' is used, where the theta parameter is estimated internally; ideal for over-dispersed events/person-time outcomes.<br>**continuous**<br>A gaussian distribution with link = 'identity' is used. |
| formula | (Optional) Default NULL. An object of class "formula" (or one that can be coerced to that class) which provides the complete model formula, similar to the formula for the `glm` function in R (e.g. 'Y ~ X + Z1 + Z2 + Z3'). Can be supplied as a character or formula object. If no formula is provided, Y and X must be provided. |
| Y | (Optional) Default NULL. Character argument which specifies the outcome variable. Can optionally provide a formula instead of Y and X variables. |
| X | (Optional) Default NULL. Character argument which specifies the exposure variable (or treatment group assignment), which can be binary, categorical, or continuous. This variable can be supplied as a factor variable (for binary or categorical exposures) or a continuous variable. For binary/categorical exposures, X should be supplied as a factor with the lowest level set to the desired referent. Numeric variables are accepted, but will be centered. Character variables are not accepted and will throw an error. Can optionally provide a formula instead of Y and X variables. |
| Z | (Optional) Default NULL. List or single character vector which specifies the names of covariates or other variables to adjust for in the `glm` function. All variables should either be factors, continuous, or coded 0/1 (i.e. not character variables). Does not allow interaction terms. |
| subgroup | (Optional) Default NULL. Character argument that indicates subgroups for stratified analysis. Effects will be reported for each category of the subgroup variable. Variable will be automatically converted to a factor if not already. |
| offset | (Optional, only applicable for rate/count outcomes) Default NULL. Character argument which specifies the person-time denominator for rate outcomes to be included as an offset in the Poisson regression model. Numeric variable should be on the linear scale; function will take natural log before including in the model. |
| rate.multiplier | (Optional, only applicable for rate/count outcomes) Default 1. Numeric variable signifying the person-time value to use in predictions; the offset variable will be set to this when predicting under the counterfactual conditions. This value should be set to the person-time denominator desired for the rate difference measure and must be inputted in the units of the original offset variable (e.g. if the offset variable is in days and the desired rate difference is the rate per 100 person-years, `rate.multiplier` should be inputted as $365.25 \times 100$). |
| exposure.scalar | (Optional, only applicable for continuous exposure) Default 1. Numeric value to scale effects with a continuous exposure. This option facilitates reporting effects for an interpretable contrast (i.e. magnitude of difference) within the continuous exposure. For example, if the continuous exposure is age in years, a multiplier of 10 would result in estimates per 10-year increase in age rather than per a 1-year increase in age. |
| exposure.center | (Optional, only applicable for continuous exposure) Default TRUE. Logical or numeric value to center a continuous exposure. This option facilitates reporting effects at the mean value of the exposure variable, and allows for a mean value to be provided directly to the function in cases where bootstrap resampling is being conducted and a standardized centering value should be used across all bootstraps. See note below on continuous exposure variables for additional details. |
| R | (Optional) Default 200. The number of data resamples to be conducted to produce the bootstrap confidence interval of the estimate. |
| clusterID | (Optional) Default NULL. Character argument which specifies the variable name for the unique identifier for clusters. This option specifies that clustering should be accounted for in the calculation of confidence intervals. The `clusterID` will be used as the level for resampling in the bootstrap procedure. |
| parallel | (Optional) Default "no." The type of parallel operation to be used. Available options (besides the default of no parallel processing) include "multicore" (not available for Windows) or "snow." This argument is passed directly to `boot`. See note about setting seeds and parallel computing. |
| ncpus | (Optional, only used if parallel is set to "multicore" or "snow") Default 1. Integer argument for the number of CPUs available for parallel processing/ number of parallel operations to be used. This argument is passed directly to `boot`. |

Most users will only need to call the `gComp` function to estimate the effects of interest. Arguments to be supplied are listed in Table 1 (and examples of how to call the function are provided below in the Results section and S1 Appendix). Users can supply individual variable names for the exposure, outcome, and covariates, or can provide a model formula. The `gComp` function (and also `pointEstimate`) does not allow for interaction terms, however subgroup analysis is possible by specifying the variable name in the dataset corresponding to the subgroup classification, which automatically adds an interaction term between the subgroup variable and the exposure to the model formula. Both functions also allow for the specification of a categorical (in addition to binary) exposure. In cases of single-level clustered data, the `gComp` function can conduct bootstrap resampling at the cluster, instead of individual sample, level by specifying the variable identifying the cluster in the `clusterID` argument.

Output of the `gComp` function is a list with several pieces of data, including parameter estimates and 95% confidence intervals for the effect measures (e.g. for a binary outcome, this would include risk difference, risk ratio, odds ratio, and number needed to treat). Confidence intervals are not reported for the number needed to treat given the primary utility of the number needed to treat for communication and the challenges in construction and interpretation of the number needed to treat confidence interval when the confidence interval for the risk difference crosses the null [24,25]. Additional output includes marginal mean predicted outcomes for each exposure level. Users can visualize the distribution of parameter estimates over all bootstrap resamples of the data by plotting the resulting data with the base R `plot()` call to the output of the `gComp` function, which provides a quantile-quantile plot [26] and histogram of all parameter estimates (see S1 Appendix).

Bootstrap resampling is necessary to estimate accurate 95% confidence intervals for the population-standardized marginal effects obtained with g-computation, since the standard errors for the coefficients from the underlying parametric model (covariate-conditional effects) are no longer applicable [16,22]. We recommend setting the number of bootstrap resamples (`R`) to 1000 for the final analysis. However, this can result in potentially long runtimes, depending on the computing power of the user’s computer (>30min). Thus, exploratory analyses can be conducted with a lower number of bootstraps (default is `R = 200`, which should compute on datasets of 5000-10000 observations in <60s).

Package code was written in R version 4.1.2 [27], and the package was built in RStudio [28] using `devtools` and `roxygen2` to generate and populate the package documentation [29,30]. `riskCommunicator` is open-source and freely available on GitHub (https://github.com/jgrembi/riskCommunicator) and Comprehensive R Archive Network (https://CRAN.R-project.org/package=riskCommunicator).

## Results

We demonstrate the utility of `riskCommunicator` using the teaching data set from the Framingham Heart Study [31], a prospective cohort study of cardiovascular disease conducted in Framingham, Massachusetts. The use of these data for the purposes of this package were approved on 11 March 2019 (request #7161) by National Institutes of Health/National Heart, Lung, and Blood Institute. These data were altered prior to receipt by the authors to ensure an anonymous dataset that protects patient confidentiality. This project was deemed by the Institutional Review Board at Emory University to not be research with human subjects and therefore did not require IRB review or consent from participants. The following analysis was conducted among 4,240 participants who conducted a baseline exam and were free of prevalent coronary heart disease when they entered the study in 1956. Participants were followed for 24 years for the combined outcome of cardiovascular disease or death due to any cause. A complete vignette highlighting the full range of analyses that are available with `riskCommunicator` is available on Comprehensive R Archive Network (CRAN).

A relatively straightforward research aim for these data would be to estimate the effect of having prevalent diabetes at the beginning of the study on the 24-year risk of cardiovascular disease or death, adjusting for the potential confounders, including patient’s age, sex, body mass index, smoking status (current smoker or not), and prevalence of hypertension. For a binary outcome, `riskCommunicator` estimates the risk difference, risk ratio, odds ratio, and number needed to treat. The output of the `gComp` function for this analysis as follows reports the strong effect of diabetes on cardiovascular disease and mortality (Table 2):

**Table 2.** Effect of prevalent diabetes at the beginning of the study on the 24-year risk of cardiovascular disease or death among 4,240 participants in the Framingham Heart Study.

|  | `riskCommunicator` | Standard regression models* |
| --- | --- | --- |
|  | Marginal effect | Covariate-conditional effect |
| Effect measure | estimate<sup>†</sup> (95% CI) | estimate<sup>†</sup> (95% CI) |
| Risk difference | 0.29 (0.20, 0.39) | N/A<sup>‡</sup> |
| Risk ratio | 1.70 (1.48, 1.97) | 1.49 (1.33, 1.66) |
| Odds ratio | 4.55 (2.77, 9.09) | 4.55 (2.66, 7.78) |
| Number needed to treat | 3.48 | N/A<sup>‡</sup> |

\*Log-linear regression for the risk difference, Poisson approximation of log-binomial regression with robust variance for the risk ratio, logistic regression for the odds ratio.
<sup>†</sup>Adjusted for patient's age, sex, body mass index (BMI), smoking status (current smoker or not), and prevalence of hypertension.
<sup>‡</sup>Log-linear model did not converge

~~~
binary.res <- gComp(data = cvdd, Y = “cvd_dth”, X =
“DIABETES”, Z = c(“AGE”, “SEX”, “BMI”, “CURSMOKE”, “PREVHYP”),
outcome.type = “binary”, R = 1000)
~~~

The absolute 24-year risk of cardiovascular disease or death due to any cause was 29% (95% CI: 20, 40) higher among subjects with diabetes at baseline compared to subjects without diabetes at baseline. The relative 24-year risk was 70% (95% CI: 48, 97) higher. Because the outcome was common (41.8%), the odds ratio (4.55) is highly inflated compared to the risk ratio (1.70). This is a clear example where the odds ratio may be misleading since the odds ratio is commonly misinterpreted as a risk ratio. Furthermore, the relative effect may be interpreted as much larger than the absolute effect, even though the absolute risk difference more closely corresponds to the expected additional number of cases due to diabetes. For public health communication, the number needed to treat derived from the risk difference (1/risk difference) provides an easily interpreted estimate of the magnitude of effect. We would expect that only 4 additional persons would need to have diabetes at baseline to observe an increase in the number of cases of cardiovascular disease or death by 1 over 24 years of follow-up.

We may also be interested in the effect of diabetes on the rate of cardiovascular disease or death, incorporating person-time at risk. If the Framingham Heart Study were an open cohort with variable follow-up time, rate-based effects would be more appropriate than risk-based measures, which assume a constant follow-up period. In addition, we may be interested in effects stratified by a potential effect measure modifier, such as participant sex. `riskCommunicator` can estimate the incidence rate difference and incidence rate ratio by sex for this analysis (Fig 1). As the person-time variable has units of days, rates are reported per 100 person-years by using the `rate.multiplier` option.

**Fig 1.**
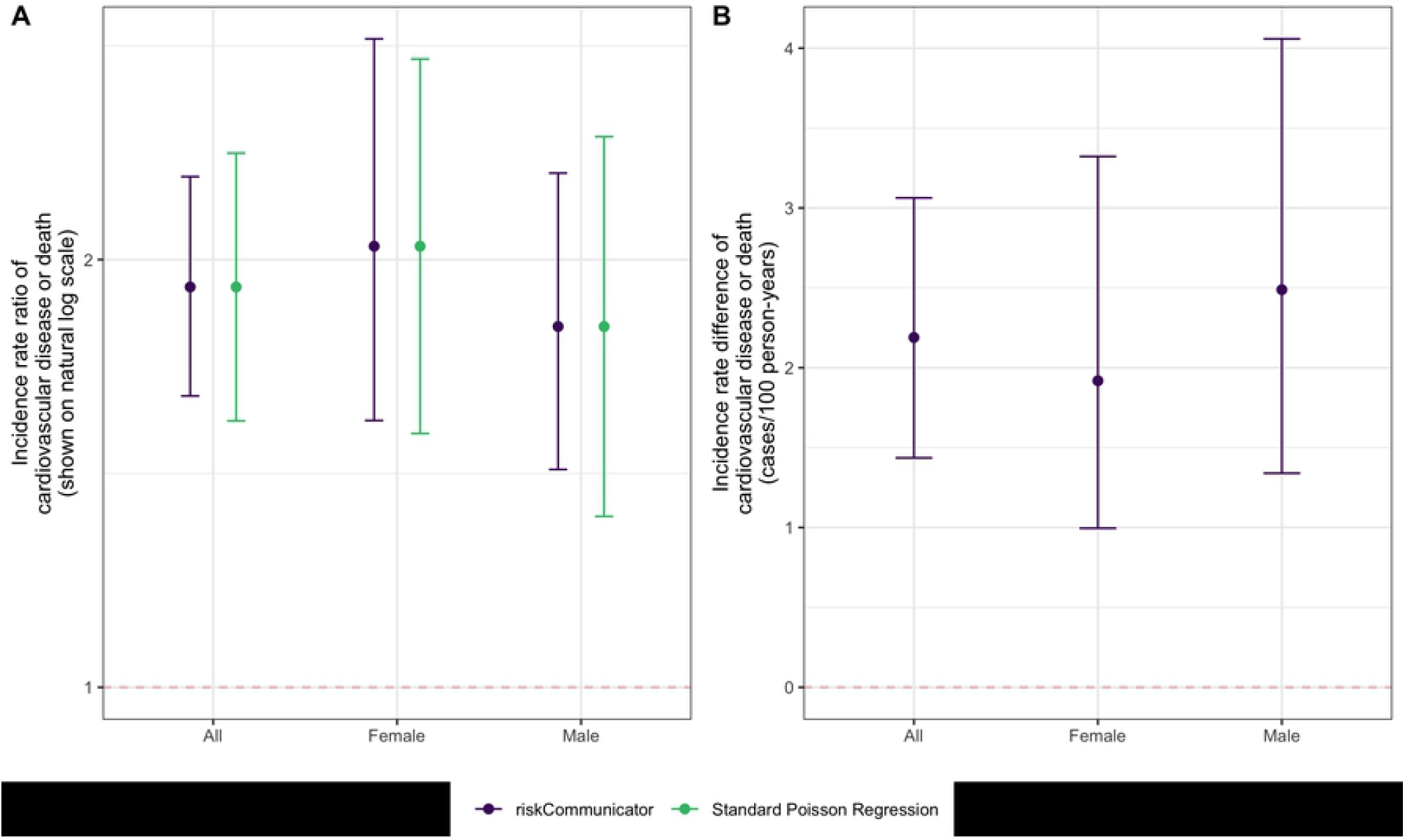
Effect of having prevalent diabetes at the beginning of the study on the 24-year risk of cardiovascular disease or death overall and stratified by sex among 4,240 participants in the Framingham Heart Study. A) Incidence rate ratio. B) Incidence rate difference. `riskCommunicator` was used to obtain marginal effect estimates (purple) and Poisson regression was used to obtain covariate-conditional estimates (green; not available for incidence rate difference). All models were adjusted for patient’s age, sex, body mass index, smoking status (current smoker or not), and prevalence of hypertension. Each point represents the point estimate and error bars show the 95% CI.

~~~
rate.res.subgroup <- gComp(data = cvdd.t, Y = “cvd_dth”, X =
“DIABETES”, Z = c(“AGE”, “SEX”, “BMI”, “CURSMOKE”, “PREVHYP”),
subgroup = “SEX”, outcome.type = “rate”, rate.multiplier =
365.25*100, offset = “timeout”, R = 1000)
~~~

There is evidence for effect modification on the additive scale. The absolute rate of cardiovascular disease or death due to any cause is 2.49 cases/100 person-years (95% CI: 1.34, 4.06) higher among males with diabetes compared to males without diabetes. In contrast, the effect among women is smaller: the absolute rate of cardiovascular disease or death is 1.92 cases/100 person-years (95% CI: 1.00, 3.32) higher among females with diabetes compared to females without diabetes. The relative effects suggest effect modification in the opposite direction on the multiplicative scale, such that the effect of diabetes is stronger among females compared to males. This difference is observed because the baseline rate of cardiovascular disease and death without diabetes is higher among males (2.77 cases/100 person-years) than females (1.64 cases/100 person-years), such that with the relative effect, the greater absolute effect among males is diluted by their higher baseline risk.

The overall incidence rate ratio in the total study population (1.91, 95% CI: 1.60, 2.29) can be estimated using the same code as above without the subgroup option. As expected, the incidence rate ratio is further from the null than the risk ratio, but closer to the null than the odds ratio (Table 1). This relationship among the magnitudes of these effect measures is expected due to their mathematical properties, and specifically the differences in the denominators of risk (total population), rates (person-time at risk), and odds (non-cases at the end of follow-up). The estimation of these effects with standard regression models is not trivial. To estimate the risk difference and risk ratio, we used log-binomial and log-linear regression, respectively. However, in these data, both models fail to converge, and the Poisson approximation with robust variance was necessary to estimate the risk ratio. The risk ratio estimate from g-computation (confidence limit ratio: 1.33) had slightly lower precision compared to the estimate from Poisson regression with robust variance (confidence limit ratio: 1.25). Minor differences in the magnitude of the estimates can be attributed to the difference between the covariate-conditional effects (as estimated by Poisson regression) and the marginal effects (as estimated by `riskCommunicator`; Table 2). Poisson regression could also be used to estimate the incidence rate ratios, resulting in equivalent magnitudes of estimates as those from `riskCommunicator`, but slightly less precision (Table 3). Adjusted incidence rate differences are not easy to estimate using standard regression models, but are readily available from `riskCommunicator`.

Finally, an additional useful output of the package is the estimation of marginal mean predicted outcomes for each exposure level. These predicted means are standardized over the observed values of covariates included in the model, and therefore are not specific to set values of the covariates. This difference is a major advantage over the usual `predict` function in R, and similar functions in other statistical programs such as the `lsmeans` statement in Statistical Analysis System (SAS), which can only predict outcomes at specific values of the other covariates.

## Conclusions

`riskCommunicator` facilitates the presentation of a wide range of effect measures with a simple user experience, similar to running a linear regression model in R. For binary outcomes, effects are modeled using logistic regression, which preserves the preferable statistical qualities usually associated with odds ratios and applies them to the estimation of risk ratios and risk differences. The package also facilitates the presentation of incidence rate differences, which are difficult to obtain with standard generalized linear models. Finally, the package supports assessment of additive effect measure modification by reporting difference effects, which is important since contradictory evidence for effect modification between the additive and multiplicative scales is common. While effect modification on the additive scale can be more relevant to public health [32,33], it is often harder to estimate with standard regression models [34,35].

It is important to highlight that the g-computation approach produces marginal rather than covariate-conditional effect estimates. In a multivariable model, the effect estimates derived directly from the covariate coefficients are covariate-conditional, interpreted as the associations given constant values of the other variables (or informally, “holding all other variables constant”) [32]. Covariate-conditional effects are difficult to interpret for non-collapsible effect measures like the odds ratio [36]. Therefore, the reporting of marginal effects, in which the effect is standardized over the covariate distribution of the total study population, may be preferable in many cases. The marginal effect is interpreted as the average treatment effect in the total population and is the primary effect of interest in randomized trials and in many observational settings where causal inference is the goal [37].

One potential limitation to the g-computation approach is the use of bootstrap for the confidence intervals. Bootstrapping is conservative compared to closed form solutions for the variance (e.g. those used to estimate Wald confidence intervals), such that the confidence intervals from bootstrapping can be slightly wider than alternatives. However, in the examples above, precision improved for the rate ratios. In addition, the precision loss is often not extreme when it occurs, and bootstrapped confidence intervals are more appropriate when the distributional assumptions or approximations of the parameter may not be valid [32]. By using percentiles of the simulated distribution of estimates from the bootstrap, one can avoid the need to calculate the standard deviation of estimates under the normal distribution assumption [38]. The use of bootstrap allows for flexibility to estimate many effects with the same framework, including allowing for clustering with bootstrap at the cluster level.

The g-computation approach can also be limited in settings with a continuous exposure variable. For example, for a binary outcome, because the underlying parametric model is logistic regression, the risks will be estimated to be linear on the log-odds (logit) scale, such that the odds ratio for any one unit increase in the continuous variable is constant. However, the risks will not be linear on the linear (risk difference) or log (risk ratio) scales, such that these parameters will not be constant across the range of the continuous exposure. The g-computation approach requires setting one specific exposure contrast within the range of the continuous exposure. Therefore, users should be aware that the risk difference, risk ratio, number needed to treat (for a binary outcome) and the incidence rate difference (for a rate/count outcome) reported do not necessarily apply across the entire range of the continuous exposure. We mitigate this issue by reporting the estimates for a relevant contrast within the exposure variable by first centering the variable at the mean and allowing users to specify a scaling factor for the contrast.

While other software packages are available to conduct more complex analyses with the g-computation approach, `riskCommunicator` has been designed to be more accessible to the average data analyst. For example, the GFORMULA macro for SAS [20] and the `gfoRmula` package in R [18] are targeted to longitudinal data with time-varying covariates. The `qgcomp` package combines g-computation with weighted quantile sum regression to estimate the effects of mixtures [39]. The `tmle3` package in R includes g-computation but is designed to enable a more comprehensive set of analyses to estimate Targeted Minimum Loss-Based Estimation (TMLE) parameters [19], which requires advanced training even for doctoral-level epidemiologists. The focus of `riskCommunicator` alternatively is on facilitating the presentation of relevant and interpretable effect measures in relatively simple time-fixed settings. The application of g-computation in these more traditional settings can help overcome the gap for less experienced users between traditional regression modeling-based methods and the g-methods, which are at the vanguard of epidemiologic methods development [40]. More importantly, `riskCommunicator` can facilitate the communication of effects of exposures and interventions and ultimately further the public health impact of epidemiologic and statistical research.

## Data Availability

The package can be downloaded from the Comprehensive R Archive Network (CRAN) version 1.0.0 at https://CRAN.R-project.org/package=riskCommunicator. A development version, issue logging, and support can be found at https://github.com/jgrembi/riskCommunicator. The package includes two vignettes, one included as Supporting Information along with this manuscript and another targeted for new R users with additional details which can be downloaded from CRAN. The dataset (?framingham?) included in the package is from The Framingham Heart Study and it?s use for the purpose of this package was approved on 11 March 2019 (request #7161) by the National Institutes of Health/National Heart, Lung, and Blood Institute. This is a teaching dataset and specific methods were employed to ensure an anonymous dataset that protects patient confidentiality.

https://CRAN.R-project.org/package=riskCommunicator

## Acknowledgements

We thank Dr. Charlie Poole for his feedback on key functions of the package. We thank Helen Pitchik and Solis Winters for beta testing and useful suggestions on formatting results for downstream plotting functionality.

## References

1. Pocock SJ, Collier TJ, Dandreo KJ, de Stavola BL, Goldman MB, Kalish LA, et al. Issues in the reporting of epidemiological studies: a survey of recent practice. BMJ. 2004;329:883. doi:10.1136/bmj.38250.571088.55

2. Vandenbroucke JP, von Elm E, Altman DG, Gøtzsche PC, Mulrow CD, Pocock SJ, et al. Strengthening the Reporting of Observational Studies in Epidemiology (STROBE): explanation and elaboration. Epidemiology. 2007;18: 805–835. doi:10.1097/EDE.0b013e3181577511

3. Knol MJ, Le Cessie S, Algra A, Vandenbroucke JP, Groenwold RHH. Overestimation of risk ratios by odds ratios in trials and cohort studies: alternatives to logistic regression. Canadian Medical Association Journal. 2012;184: 895–899. doi:10.1503/cmaj.101715

4. Holcomb WL, Chaiworapongsa T, Luke DA, Burgdorf KD. An odd measure of risk: use and misuse of the odds ratio. Obstet Gynecol. 2001;98: 685–688. doi:10.1016/s0029-7844(01)01488-0

5. Persoskie A, Ferrer RA. A Most Odd Ratio. Am J Prev Med. 2017;52: 224–228. doi:10.1016/j.amepre.2016.07.030

6. Katz KA. The (Relative) Risks of Using Odds Ratios. Arch Dermatol. 2006;142: 761–764. doi:10.1001/archderm.142.6.761

7. Schwartz LM, Woloshin S, Dvorin EL, Welch HG. Ratio measures in leading medical journals: structured review of accessibility of underlying absolute risks. BMJ. 2006;333: 1248. doi:10.1136/bmj.38985.564317.7C

8. Noordzij M, van Diepen M, Caskey FC, Jager KJ. Relative risk versus absolute risk: one cannot be interpreted without the other. Nephrol Dial Transplant. 2017;32: ii13–ii18. doi:10.1093/ndt/gfw465

9. Zou G. A modified poisson regression approach to prospective studies with binary data. Am J Epidemiol. 2004;159: 702–706.

10. Greenland S. Estimating standardized parameters from generalized linear models. Stat Med. 1991;10: 1069–1074. doi:10.1002/sim.4780100707

11. Kovalchik SA, Varadhan R, Fetterman B, Poitras NE, Wacholder S, Katki HA. A general binomial regression model to estimate standardized risk differences from binary response data. Stat Med. 2013;32: 808–821. doi:10.1002/sim.5553

12. Xu Y, Cheung YB, Lam KF, Tan SH, Milligan P. A Simple Approach to the Estimation of Incidence Rate Difference. Am J Epidemiol. 2010;172: 334–343. doi:10.1093/aje/kwq099

13. Yelland LN, Salter AB, Ryan P. Relative Risk Estimation in Randomized Controlled Trials: A Comparison of Methods for Independent Observations. The International Journal of Biostatistics. 2011;7: 1–31. doi:10.2202/1557-4679.1278

14. Localio AR, Margolis DJ, Berlin JA. Relative risks and confidence intervals were easily computed indirectly from multivariable logistic regression. J Clin Epidemiol. 2007;60: 874–882. doi:10.1016/j.jclinepi.2006.12.001

15. Bieler GS, Brown GG, Williams RL, Brogan DJ. Estimating model-adjusted risks, risk differences, and risk ratios from complex survey data. Am J Epidemiol. 2010;171: 618–623. doi:10.1093/aje/kwp440

16. Ahern J, Hubbard A, Galea S. Estimating the effects of potential public health interventions on population disease burden: a step-by-step illustration of causal inference methods. Am J Epidemiol. 2009;169: 1140–1147. doi:10.1093/aje/kwp015

17. Westreich D, Cole SR, Young JG, Palella F, Tien PC, Kingsley L, et al. The parametric g-formula to estimate the effect of highly active antiretroviral therapy on incident AIDS or death. Stat Med. 2012;31: 2000–2009. doi:10.1002/sim.5316

18. Lin V, McGrath S, Zhang Z, Logan RW, Petito LC, Young JC, et al. gfoRmula: Parametric G-formula. 30 Jan 2020 [cited 7 Feb 2020]. Available: https://cran.r-project.org/web/packages/gfoRmula/index.html

19. Coyle J. R/tmle: The Extensible TMLE framework. [cited 7 Feb 2020]. Available: https://tlverse.org/tmle3/

20. Harvard Program on Causal Inference: Software. [cited 7 Feb 2020]. Available: https://www.hsph.harvard.edu/causal/software/

21. Robins J. A new approach to causal inference in mortality studies with a sustained exposure period—application to control of the healthy worker survivor effect. Mathematical Modelling. 1986;7: 1393–1512. doi:10.1016/0270-0255(86)90088-6

22. Snowden JM, Rose S, Mortimer KM. Implementation of G-computation on a simulated data set: demonstration of a causal inference technique. Am J Epidemiol. 2011;173: 731–738. doi:10.1093/aje/kwq472

23. Efron B, Tibshirani R. Bootstrap Methods for Standard Errors, Confidence Intervals, and Other Measures of Statistical Accuracy. Statistical Science. 1986;1: 54–75.

24. Hutton JL. Number needed to treat: properties and problems. Journal of the Royal Statistical Society: Series A (Statistics in Society). 2000;163: 381–402. doi:10.1111/1467-985X.00175

25. Stang A, Poole C, Bender R. Common problems related to the use of number needed to treat. Journal of Clinical Epidemiology. 2010;63: 820–825. doi:10.1016/j.jclinepi.2009.08.006

26. Wilk MB, Gnanadesikan R. Probability Plotting Methods for the Analysis of Data. Biometrika. 1968;55: 1–17. doi:10.2307/2334448

27. R Core Team. R: A language and environment for statistical computing. Vienna, Austria: R Foundation for Statistical Computing; 2013. Available: http://www.R-project.org/

28. RStudio Team. RStudio: Integrated Development for R. tBoston, MA: RStudio, Inc.; 2015. Available: http://www.rstudio.com/

29. Wickham H, Chang W. devtools: Tools to Make Developing R Packages Easier. 2017. Available: https://CRAN.R-project.org/package=devtools

30. Wickham H, Danenberg P, Eugster M. roxygen2: In-Line Documentation for R. Available: https://CRAN.R-project.org/package=roxygen2

31. National Heart, Lunch, and Blood Institute. Teaching Datasets. [cited 11 Mar 2019]. Available: https://biolincc.nhlbi.nih.gov/teaching/

32. Rothman KJ, Greenland S, Lash TL. Modern Epidemiology. 3rd ed. Philadelphia, PA: Lippincott William & Wilkins; 2008.

33. VanderWeele TJ. Explanation in Causal Inference: Methods for Mediation and Interaction. New York, NY: Oxford University Press; 2015.

34. Hosmer DW, Lemeshow S. Confidence Interval Estimation of Interaction. Epidemiology. 1992;3: 452.

35. Richardson DB, Kaufman JS. Estimation of the Relative Excess Risk Due to Interaction and Associated Confidence Bounds. Am J Epidemiol. 2009;169: 756–760. doi:10.1093/aje/kwn411

36. Pang M, Kaufman JS, Platt RW. Mixing of confounding and non-collapsibility: a notable deficiency of the odds ratio. Am J Cardiol. 2013;111: 302–303. doi:10.1016/j.amjcard.2012.09.002

37. Funk MJ, Westreich D, Wiesen C, Stürmer T, Brookhart MA, Davidian M. Doubly Robust Estimation of Causal Effects. Am J Epidemiol. 2011;173: 761–767. doi:10.1093/aje/kwq439

38. Greenland S. Interval estimation by simulation as an alternative to and extension of confidence intervals. Int J Epidemiol. 2004;33: 1389–1397. doi:10.1093/ije/dyh276

39. Keil Alexander P., Buckley Jessie P., O’Brien Katie M., Ferguson Kelly K., Zhao Shanshan, White Alexandra J. A Quantile-Based g-Computation Approach to Addressing the Effects of Exposure Mixtures. Environmental Health Perspectives. 128: 047004. doi:10.1289/EHP5838

40. Naimi AI, Cole SR, Kennedy EH. An introduction to g methods. Int J Epidemiol. 2017;46: 756–762. doi:10.1093/ije/dyw323

